# A common variant that alters SUN1 degradation associates with hepatic steatosis and metabolic traits in multiple cohorts

**DOI:** 10.1101/2022.10.19.22280653

**Authors:** Kapil K. Upadhyay, Xiaomeng Du, Yanhua Chen, Raymond Zhao, Elizabeth K. Speliotes, Graham F. Brady

## Abstract

Nonalcoholic fatty liver (NAFL) and nonalcoholic steatohepatitis (NASH) represent a genetically and phenotypically diverse entity with no approved therapy, making it imperative to define the spectrum of pathways contributing to its pathogenesis. Rare variants in genes encoding nuclear envelope proteins cause lipodystrophy that includes early-onset NASH; we hypothesized that common variants in nuclear envelope-related genes might also contribute to hepatic steatosis and NASH. In an association meta-analysis of nuclear envelope-related coding variants in three large cohorts (N>120,000 participants), rs6461378 (*SUN1* H118Y) was the top steatosis-associated variant (*P*<0.001). In ancestrally distinct validation cohorts, rs6461378 positively associated with NASH-related metabolic traits including increased serum fatty acids, decreased HDL, type 2 diabetes, hypertension, and cardiovascular disease. SUN1 H118Y was subject to increased proteasomal degradation relative to wild-type SUN1 in Huh7 cells, and SUN1 H118Y-expressing cells exhibited insulin resistance and increased lipid accumulation. Collectively, these data support a potential causal role for rs6461378 in NASH and metabolic disease.

**Lay Summary:** A common genetic variant that leads to an amino acid change in the nuclear envelope protein SUN1 was found to positively associate with hepatic steatosis in a meta-analysis of genomic data from multiple large cohorts. Follow-up studies in separate validation cohorts demonstrated strong positive associations with metabolic traits that are linked to nonalcoholic fatty liver disease, including insulin resistance, type 2 diabetes, hypertension, and cardiovascular disease. Testing of this variant in cell culture demonstrated biochemical differences from wild-type SUN1, with increased proteasomal degradation of the H118Y variant, decreased sensitivity to insulin, and increased lipid accumulation, suggesting that this is a functional variant with a potential causal role in human disease.

## Introduction

Nonalcoholic fatty liver disease (NAFLD) and nonalcoholic steatohepatitis (NASH) represent a genetically and phenotypically diverse disease entity that is now the most common liver disease in many countries and has no approved medical therapy (Cotter and Rinella, 2020; Konerman et al., 2018; Paik et al., 2020; Wong et al., 2015; Younossi et al., 2018). Therefore, it is imperative to define the full spectrum of pathways that contribute to its pathogenesis to allow development of effective treatments and identification of the patients who are most likely to benefit from those treatments.

Rare variants in genes encoding nuclear envelope proteins, particularly *LMNA*, encoding A-type lamin proteins, cause lipodystrophy syndromes that include early-onset insulin resistance and NASH (Ajluni et al., 2017; Brady et al., 2018a; Shackleton et al., 2000). Genetic animal models support the pathogenicity of these rare variants and have pointed to putative mechanisms by which nuclear envelope dysfunction might contribute to disease in humans (Kwan et al., 2017; Le Dour et al., 2017; Shin et al., 2019; Wojtanik et al., 2009). However, it has been unclear whether additional variants in nuclear envelope-related genes might contribute to NAFLD/NASH and associated metabolic diseases in the population at large. In a small cohort of twins and siblings, we found that variants in some of these genes clustered among participants with NAFLD (Brady et al., 2018b). We therefore hypothesized that additional, and perhaps more common, variants in nuclear envelope-related genes might also contribute to hepatic steatosis and NAFLD at a population level.

Multiple genome-wide association studies (GWAS) have demonstrated associations between NAFLD and several loci including those containing the *PNPLA3, TM6SF2, GCKR*, and *MBOAT7* genes (Anstee et al., 2016; Speliotes et al., 2011). However, the estimated heritability of NAFLD based on twin and familial studies may approach 50%, which is not explained by currently known loci, suggesting more NAFLD-associated loci likely exist but have yet to be identified (Anstee et al., 2016; Cui et al., 2016; Grove et al., 2016; Loomba et al., 2015).

To address the possibility that nuclear envelope-related variants might contribute to NAFLD at a population level, we employed a meta-analysis of GWAS data derived from three large western datasets – UK Biobank (UKBB), Michigan Genomics Initiative (MGI), and the Genetics of Obesity-related Liver Disease Consortium (GOLDc) which consists of multiple well-phenotyped cohorts based in Europe and the United States (Speliotes et al., 2011; Palmer et al., 2021). Here we report the identification of a common coding variant in the nuclear envelope-related gene *SUN1* which positively associated with steatosis in all three discovery cohorts, as well as with related metabolic traits in multiple validation cohorts; moreover, the encoded protein variant demonstrated a biochemical phenotype relative to wild-type (WT) SUN1 when tested *in vitro*, suggesting a potentially direct functional impact of this variant in metabolic disease in humans.

## Materials and Methods

### Ethics Statement

Where relevant, all research in this study was approved by the Institutional Review Board (IRB) of the University of Michigan (Ann Arbor, MI). UKBB protocols were approved by the National Research Ethics Service Committee, and all participants provided written informed consent. Analyses of UKBB data in this project were conducted under UK Biobank Resource Project 18120. GOLDc participants provided written informed consent upon enrollment, and study protocols for each component cohort were approved by the relevant site-specific IRB (Palmer et al., 2021). IRB approval was not required to use BioBank Japan, FinnGen, or FinMetSeq data as they are publicly available. Michigan Genomics Initiative (MGI) participants provided written informed consent approved by the University of Michigan IRB. ***Association meta-analysis***. Genomic data for analysis were derived from the following cohorts: Michigan Genomics Initiative (MGI; N=57,022), UK Biobank (UKBB; N=43,293), GOLD Consortium (N=23,521). All-ancestry genome-wide association study (GWAS) of autosomal variants was first performed for each individual dataset in SAIGE version 0.29, with hepatic steatosis as the outcome, in an additive genetic model, controlling for age, age^2^, sex, and the first 10 principal components. For the GOLDc dataset, GWAS was first performed in each of the nine component cohorts separately; sensitivity analyses by sex, study, and ancestry did not show significant heterogeneity, allowing us to combine the GOLDc data for all individuals with genetic data (N= 23,521). For UKBB and GOLDc, hepatic steatosis was determined by cross-sectional imaging (MRI and CT, respectively; Palmer et al., 2021); for MGI, the presence of hepatic steatosis was determined via natural language processing of pathology and radiology reports (Chen et al., 2021; **Table 1**). Then, all protein-coding variants (defined as missense, nonsense, or short insertion/deletion variants predicted to alter the encoded protein sequence) within 17 nuclear envelope-related genes (**Table 2)** that passed quality control (QC) in each individual cohort, including imputation score >0.85, were included in the multi-cohort meta-analysis performed in METAL (08/28/2018 release; Chen et al., 2021). Potential significance of a variant’s association with hepatic steatosis was determined after correction for multiple comparisons via the Benjamini-Hochberg method with a false-discovery rate of 0.05. Variant annotation as potentially deleterious or damaging was performed via SIFT and PolyPhen-2.

**Table 1.** Discovery cohorts and outcomes. MRI-PDFF, magnetic resonance imaging proton density fat fraction.

| **Cohort** | **Number of subjects** | **Outcome** | **Criteria for outcome** |
| --- | --- | --- | --- |
| Michigan Genomics Initiative (MGI) | 57,022 | Hepatic steatosis | Natural language processing of radiology and pathology reports (dichotomous) |
| UK Biobank (UKBB) | 43,293 | Hepatic steatosis | MRI-PDFF |
| Genetics of Obesity-related Liver Disease Consortium (GOLDc) | 23,521 | Hepatic steatosis | CT liver attenuation (Speliotes et al., 2011; Palmer et al., 2021) |

**Table 2.** Candidate genes included in genomic analysis.

| Gene | Encoded protein | Number of coding variants with data from >1 cohort | Number of coding variants with data from all cohorts |
| --- | --- | --- | --- |
| *LMNA* | A-type lamin | 2 | 0 |
| *LMNB1* | B-type lamin | 1 | 1 |
| *LMNB2* | B-type lamin | 2 | 0 |
| *TOR1AIP1* | Transmembrane protein (inner nuclear membrane) | 5 | 3 |
| *TOR1AIP2* | Transmembrane protein (ER membrane) | 1 | 1 |
| *LBR* | Transmembrane protein (inner nuclear membrane) | 7 | 5 |
| *SUN1* | Transmembrane protein (inner nuclear membrane) | 13 | 10 |
| *SUN2* | Transmembrane protein (inner nuclear membrane) | 12 | 9 |
| *TMEM43* | Transmembrane protein (inner nuclear membrane) | 8 | 4 |
| *ZMPSTE24* | Lamin-processing enzyme | 0 | 0 |
| *ICMT* | Lamin-processing enzyme | 2 | 1 |
| *FNTA* | Lamin-processing enzyme | 1 | 0 |
| *FNTB* | Lamin-processing enzyme | 1 | 0 |
| *TMPO* | Lamin binding partner | 14 | 11 |
| *BANF1* | Lamin binding partner | 0 | 0 |
| *TOR1A* | Torsin protein, in intermembrane space | 1 | 1 |
| *TOR1B* | Torsin protein, in intermembrane space | 0 | 0 |

### Phenome-wide association study (PheWAS)

Additional phenotype associations of the top steatosis-associated variant, rs6461378 (g.842031C>T; *SUN1* H118Y), were tested in separate validation cohorts – FinnGen Release 7 (N=309,154), FinMetSeq (N=19,289), BioBank Japan (updated March 12, 2022; N=260,000) – using publicly available web-based data browsers.

### Cell culture and treatment conditions

Huh7 cells were maintained in Dulbecco’s modification of Eagle’s medium (DMEM) with glucose, without L-glutamine (Lonza, Williamsport, PA) supplemented with 10% heat-inactivated fetal bovine serum (Thermo Fisher A3840202, USA) at 37°C, 5% CO2. For transfections, cells were plated at ∼60-70% confluency one day prior to transfection. Where indicated, cells were cultured in incomplete medium without serum. MG132 (Sigma-Aldrich, St. Louis, MO) was used for inhibition of the proteasome, or bafilomycin A1 (Sigma-Aldrich) for inhibition of autophagy, prior to cell lysis where indicated. Recombinant human insulin was obtained from Sigma-Aldrich (catalog number 91077C). For lipid staining, Huh7 cells were plated in 4 well chamber slides in DMEM with 10% FBS, then transfected with SUN1-mCherry the next day. After 24 hours, culture medium was changed to DMEM containing 1% fatty acid–free bovine serum albumin (Sigma-Aldrich) and oleic acid (100 µM; Sigma-Aldrich) or vehicle and incubated for 24 hours as previously described (Brady et al., 2018b). For staining, cells were fixed in 4% paraformaldehyde (PFA) in PBS for 15 mins at 22°C, then washed in PBS; neutral lipids were stained with 10 μM boron dipyrromethene 493/503 (Thermo Fisher), and quantification of green fluorescence was performed using ImageJ version 1.4.3.67.

### Plasmids and transfection

A plasmid containing the human *SUN1* transcript 1 open reading frame was purchased from Origene (RC226167; Rockville, MD). An in-frame carboxy-terminal mCherry or myc-DDK tag was inserted via NEB HiFi cloning kit (New England Biolabs, Ipswich, MA). The H118Y variant of SUN1 was generated using the QuikChange II site-directed mutagenesis kit (Agilent Technologies, Santa Clara, CA). Lipofectamine-2000 (Life Technologies, Carlsbad, CA) was used for transfection according to the manufacturer’s instructions.

### Immunofluorescence

Two days after transfection, cells were washed, then fixed with 4% PFA in PBS for 15min. Following fixation and permeabilization (0.1% Triton X-100 in PBS, 10min), washing then blocking, primary antibodies (**Supplementary Table 1**) were added (overnight, 4° C). After washing, fluorescently tagged secondary antibody (Alexa Fluor® 680; Thermo Fisher) was added (1h, 22° C), followed by DNA staining using Prolong Gold Anti-fade reagent with DAPI (Thermo Fisher). Stained cells were visualized with a Leica DM 5000B microscope, and images were acquired with a 20× objective.

### Gel electrophoresis and immunoblotting

Cells were lysed and proteins solubilized in RIPA lysis buffer (Thermo Fisher) containing protease inhibitor cocktail (Sigma-Aldrich); for phosphoprotein analysis, lysis buffer also included phosphatase inhibitor cocktail (Thermo Fisher). After centrifugation (12,000x*g*, 10min, 4° C), the protein concentration was determined (bicinchoninic acid assay, Thermo Fisher). Proteins were resolved using 4-12% Novex Tris-Glycine gels (Invitrogen) after boiling in the presence of SDS and reducing agent (2% β-mercaptoethanol). After resolution via SDS-PAGE, proteins were transferred to polyvinylidene fluoride membranes (Bio-Rad, Hercules, CA) for immunoblotting. Where indicated, data were quantified by densitometry using ImageJ version 1.4.3.67.

### Quantitative real-time polymerase chain reaction (qPCR) analysis

Total RNA from transfected Huh7 cells was isolated using RNeasy Mini Kit (Qiagen), and cDNA was synthesized using iScript cDNA Synthesis kit (Bio-Rad). Transcript levels of genes of interest were quantified using a Mastercycler ep *realplex*^*2*^ Gradient Thermocycler (Eppendorf); qPCR primer sequences are shown in **Supplementary Table 2**. Relative expression was determined after normalizing to 18S RNA and use of 2^−ΔΔCT^ method.

### Statistical analysis

Unless otherwise specified, for continuous data the two-tailed Student’s *t*-test or the Mann-Whitney U test was used to assess statistical significance, and analyses were performed using GraphPad Prism version 8. For categorical data, a two-tailed Fisher’s exact test was used. For insulin dose-response curves, nonlinear regression analysis was performed in GraphPad Prism version 8 to determine best-fit curves and statistical significance at a threshold of *P*<0.05. Where indicated, correction for multiple comparisons was performed via the Benjamini-Hochberg method with a false-discovery rate (FDR) of 0.05.

## Results

### GWAS meta-analysis identifies positive association of rs6461378 with hepatic steatosis

Potential association with hepatic steatosis was tested for 70 protein-coding variants (missense, nonsense, or short insertion/deletion variants predicted to alter the encoded protein sequence) in 17 nuclear envelope-related genes via meta-analysis of GWAS data from MGI, UKBB, and GOLDc (**Table 1, Figure 1A**); genes tested included those encoding lamins, lamin-processing enzymes, and lamin-binding proteins, including integral proteins of the inner nuclear membrane (INM; **Table 2**). Among 70 variants for which genotype data were available from at least two of three cohorts (**Supplementary Table 3**), only seven variants positively associated with hepatic steatosis in all cohorts, of which three were in *SUN1*, including two of the overall top three variants (**Table 3**). Of these, the top variant was rs6461378 (g.842031C>T; *SUN1* H118Y), which positively associated with hepatic steatosis in all cohorts (*P*<0.001) and remained significantly associated after Benjamini-Hochberg adjustment with FDR of 0.05 or via Bonferroni correction (**Figure 1B**); therefore, rs6461378 met criteria for further testing.

**Fig. 1.**
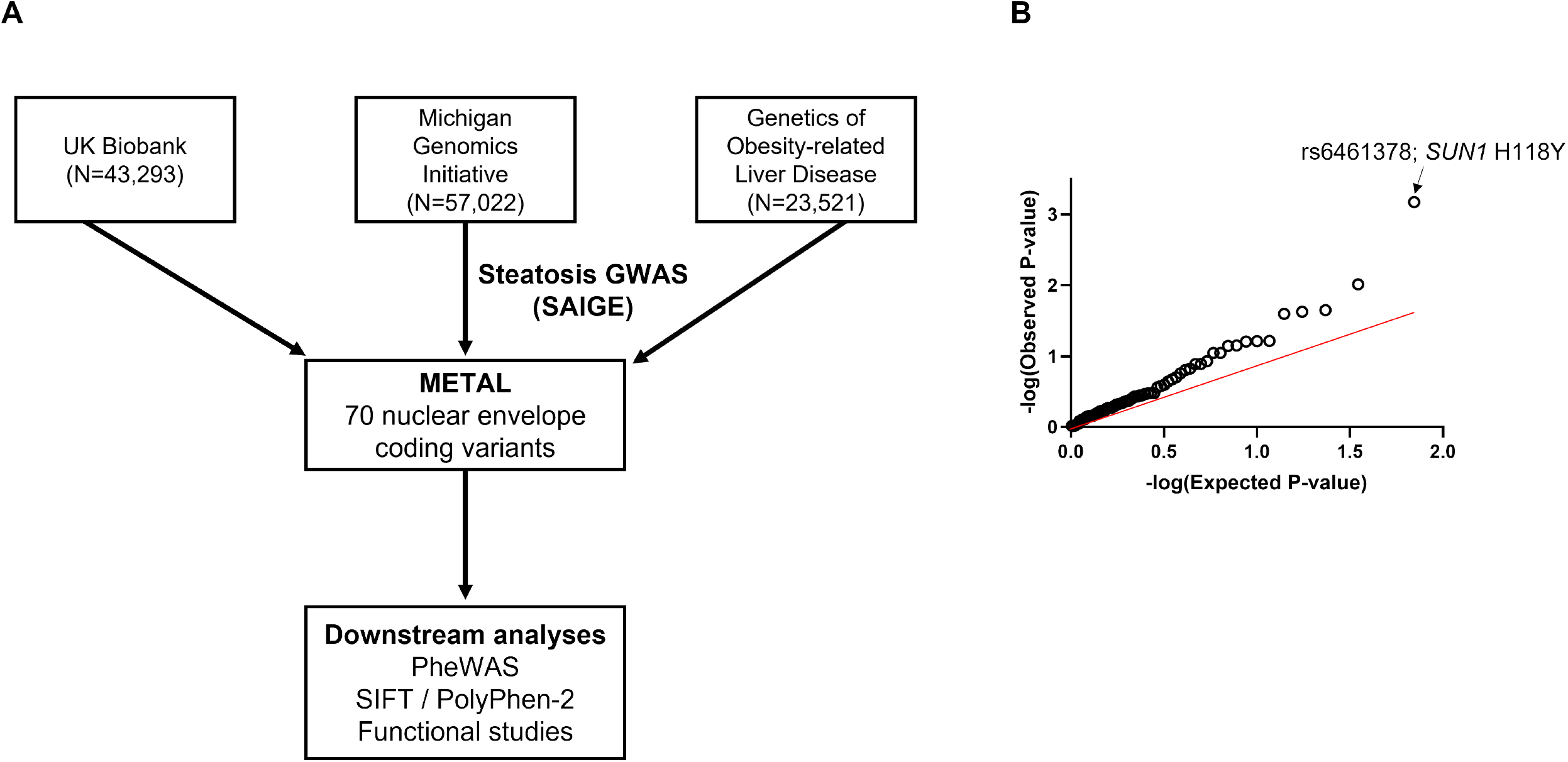
Overall study workflow and quantile–quantile plot of the hepatic steatosis association meta-analysis. (A) GWAS data from the discovery cohorts, derived in SAIGE, were subjected to meta-analysis in METAL, followed by validation studies of the top steatosis-associated variant via PheWAS, *in silico* prediction, and functional testing in cultured cells. (B) Observed versus expected -log10(*P*-value) is plotted for the 70 coding variants in nuclear envelope-related genes included in the meta-analysis; rs6461378 was the top steatosis-associated variant, as indicated.

**Table 3.**
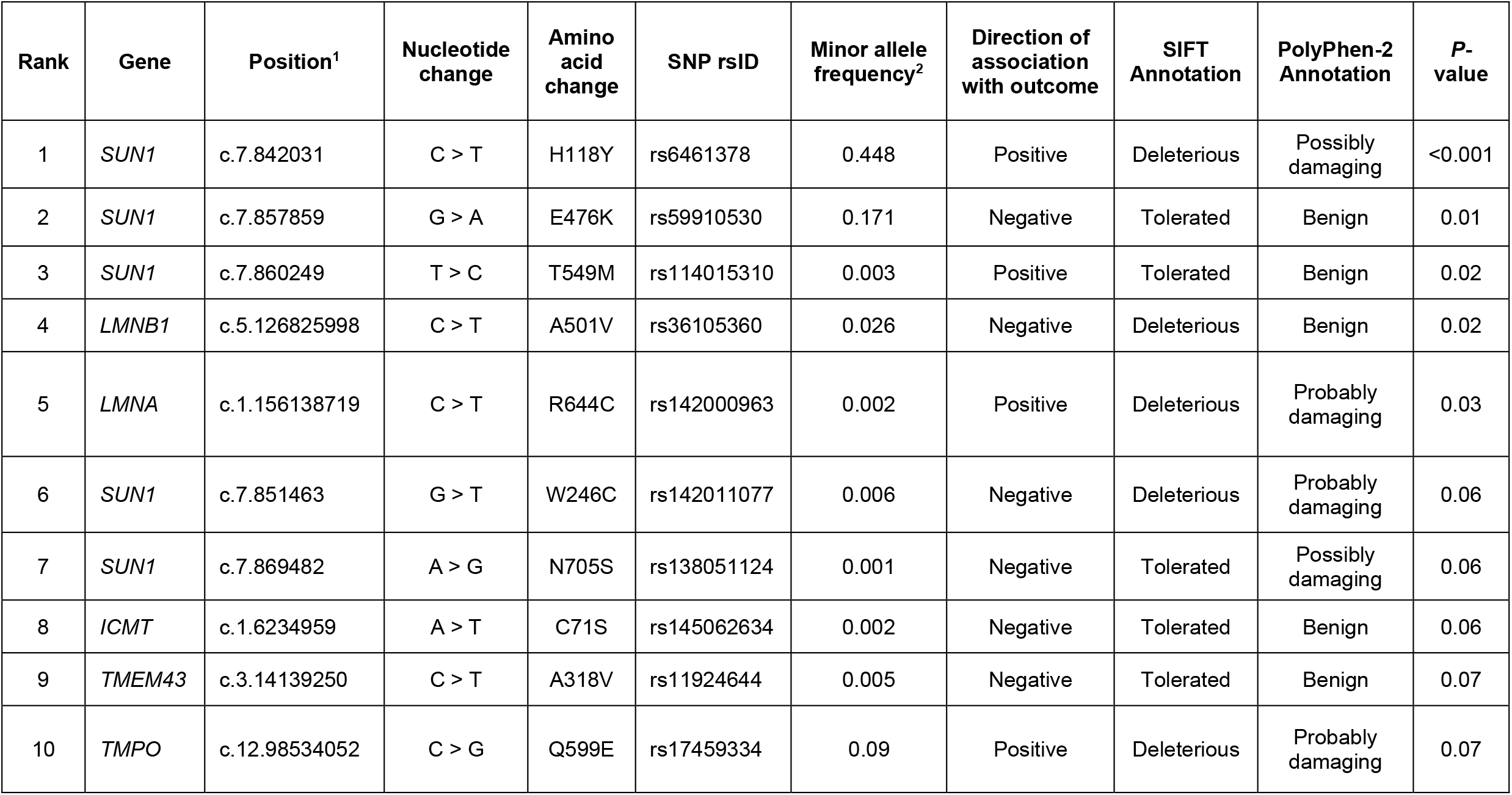
Top steatosis-associated variants in meta-analysis of discovery cohorts. ^1^, SNP location according to GRCh38.p13; ^2^, minor allele frequency in 3-cohort meta-analysis.

| Rank | Gene | Position<sup>1</sup> | Nucleotide change | Amino acid change | SNP rsID | Minor allele frequency<sup>2</sup> | Direction of association with outcome | SIFT Annotation | PolyPhen-2 Annotation | P-value |
| --- | --- | --- | --- | --- | --- | --- | --- | --- | --- | --- |
| 1 | *SUN1* | c.7.842031 | C > T | H118Y | rs6461378 | 0.448 | Positive | Deleterious | Possibly damaging | <0.001 |
| 2 | *SUN1* | c.7.857859 | G > A | E476K | rs59910530 | 0.171 | Negative | Tolerated | Benign | 0.01 |
| 3 | *SUN1* | c.7.860249 | T > C | T549M | rs114015310 | 0.003 | Positive | Tolerated | Benign | 0.02 |
| 4 | *LMNB1* | c.5.126825998 | C > T | A501V | rs36105360 | 0.026 | Negative | Deleterious | Benign | 0.02 |
| 5 | *LMNA* | c.1.156138719 | C > T | R644C | rs142000963 | 0.002 | Positive | Deleterious | Probably damaging | 0.03 |
| 6 | *SUN1* | c.7.851463 | G > T | W246C | rs142011077 | 0.006 | Negative | Deleterious | Probably damaging | 0.06 |
| 7 | *SUN1* | c.7.869482 | A > G | N705S | rs138051124 | 0.001 | Negative | Tolerated | Possibly damaging | 0.06 |
| 8 | *ICMT* | c.1.6234959 | A > T | C71S | rs145062634 | 0.002 | Negative | Tolerated | Benign | 0.06 |
| 9 | *TMEM43* | c.3.14139250 | C > T | A318V | rs11924644 | 0.005 | Negative | Tolerated | Benign | 0.07 |
| 10 | *TMPO* | c.12.98534052 | C > G | Q599E | rs17459334 | 0.09 | Positive | Deleterious | Probably damaging | 0.07 |

### In silico *analysis of rs6461378 predicts potential pathogenicity*

The identified *SUN1* H118Y variant, hereafter designated rs6461378-T, is common, with a minor allele frequency (MAF) of 0.43 in UKBB. However, it has not previously been associated with any human disease or tested for functional effect(s) in any published report. The protein encoded by *SUN1* is a ubiquitous integral protein of the INM of nucleated metazoan cells, and its function is conserved in animals and plants (Graumann et al., 2010). It is a component of the Linker of Nucleoskeleton and Cytoskeleton (LINC) complex and is known to bind to lamins at the nucleoplasmic side of the INM (Haque et al., 2006). Notably, the residue predicted to be impacted by this variant, His118 in the major isoform of SUN1, is located on the nucleoplasmic side of the INM, is well-conserved among mammals (**Supplementary Figure 1**), and is within the lamin-binding portion of the protein (Haque et al., 2006). Therefore, given the known role of lamin A/C in metabolic disease and NASH, we reasoned that rs6461378-T could plausibly play a functional role in disease. To test this possibility, we examined its potential effect on protein function *in silico* using two validated algorithms: SIFT and PolyPhen-2 (**Table 3**). The SIFT algorithm (http://sift.bii.a-star.edu.sg) for rs6461378-T produced a SIFT score of 0.03 and a prediction of “deleterious” for the primary *SUN1* transcript (transcript variant 1, encoding the major SUN1 isoform). The PolyPhen-2 algorithm (http://genetics.bwh.harvard.edu/pph2/) returned a prediction of “possibly damaging” using either the HumDiv (score = 0.91) or the HumVar (score = 0.56) model. Thus, *in silico* prediction supported a potential pathogenic role for this variant.

### Phenome-wide association study (PheWAS) of rs6461378-T confirms positive associations with NASH-related metabolic traits

Given that NAFLD is tightly linked epidemiologically to other metabolic traits including insulin resistance, type 2 diabetes, metabolic syndrome, and cardiovascular disease, we reasoned that rs6461378-T might be associated with additional NAFLD-associated traits. To test this possibility, we selected three ancestrally distinct validation cohorts for which summary statistics are publicly available: BioBank Japan (BBJ; N=260,000), FinnGen (N=309,154), and FinMetSeq (N=19,289). PheWAS using BBJ case-control data showed positive associations of rs6461378-T with type 2 diabetes (*P*=0.004), unstable angina (*P*<0.001), and ischemic stroke (*P*=0.02). Among quantitative traits, this variant strongly associated with higher systolic blood pressure (*P*<0.001), higher body mass index (*P*=0.01), and lower HDL cholesterol (*P*=0.015); see **Table 4**. Within FinnGen, the strongest positive associations were with hypertension (*P*<10^−6^) and cardiovascular disease (*P*<0.001), and an additional positive association was noted with polycystic ovarian syndrome (PCOS; *P*<0.001), which is tightly linked to insulin resistance.

**Table 4.** PheWAS associations of rs6461378-T, *SUN1* H118Y.

| Cohort | Number of participants | Phenotype | Direction of association with outcome | Beta | P-value |
| --- | --- | --- | --- | --- | --- |
| FinMetSeq | 19,289 | Postprandial serum fatty acids | Positive | 0.051 | <0.001 |
| FinnGen | 309,154 | Cardiovascular disease | Positive | 0.021 | <0.001 |
|  |  | Hypertension | Positive | 0.025 | <0.001 |
|  |  | PCOS | Positive | 0.150 | <0.001 |
| BioBank Japan | 260,000 | Type 2 diabetes | Positive | 0.027 | 0.004 |
|  |  | Unstable angina | Positive | 0.075 | <0.001 |
|  |  | HDL | Negative | -0.012 | 0.015 |
|  |  | Systolic BP | Positive | 0.011 | <0.001 |
|  |  | BMI | Positive | 0.008 | 0.010 |
|  |  | Cholelithiasis | Positive | 0.058 | <0.001 |

The positive associations of rs6461378-T with cardiovascular disease, hypertension, and elements of the metabolic syndrome in these two ancestrally distinct validation cohorts led us to explore potential associations with quantitative traits in the FinMetSeq cohort. Within this metabolically phenotyped cohort, rs6461378-T strongly associated with 2-hour postprandial serum fatty acids (*P*<0.001; **Figure 2A**), a trait that is tightly linked to insulin resistance (Axelsen et al., 1999; Wang et al., 2018). This strong positive association was unexpected, as this was one of few metabolic traits in FinMetSeq with no variants that associated with genome-wide significance (Locke et al., 2019). Indeed, Manhattan plot of GWAS data for this trait in FinMetSeq revealed a surprising finding, with single-gene peaks limited to chromosome 19 (*INSR*, encoding the insulin receptor), chromosome 5 (*OXCT1*, encoding succinyl-CoA:3-ketoacid coenzyme A transferase 1, a critical enzyme in ketone body metabolism), and chromosome 7 (*SUN1*); **Figure 2B**. Among *SUN1* variants, rs6461378-T was the top associated coding variant and was found to be in linkage disequilibrium with all positively associated non-coding *SUN1* variants with lower *P*-values (determined via LDProxy, http://ldlink.nci.nih.gov). Taken together, these data support a plausible causal link between rs6461378-T and metabolic disease, including insulin resistance and NAFLD.

**Fig. 2.**
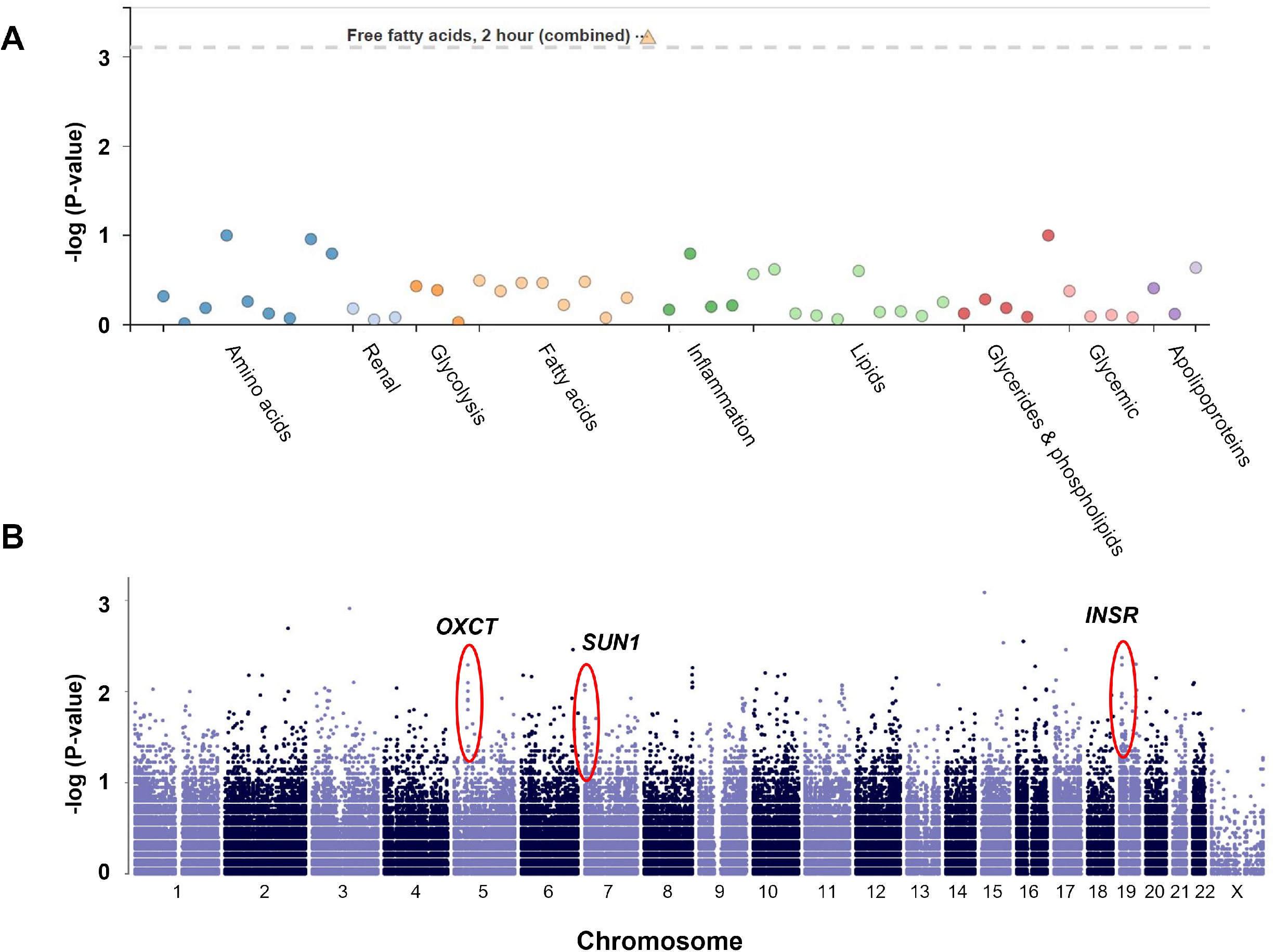
PheWAS of rs6461378-T in FinMetSeq shows strong positive association with postprandial serum fatty acids. (A) PheWAS of publicly available data from FinMetSeq study shows strong positive association with the steatosis-associated rs6461378-T and serum free fatty acids 2 hours after a test meal; dotted line represents Bonferroni-corrected significance threshold after adjustment for multiple measured metabolites. (B) Manhattan plot of GWAS data from the FinMetSeq study using postprandial serum fatty acids shows a strong single-gene peak on chromosome 7 corresponding to *SUN1*.

### SUN1 H118Y is subject to increased proteasomal degradation

Given that rs6461378-T (*SUN1* H118Y) positively associated with NAFLD and multiple related metabolic traits in our discovery and validation cohorts, and that *in silico* analysis suggested a deleterious impact on SUN1 function, we sought to test this directly in human cells. We selected Huh7 hepatoma cells for analysis as they are a robust and commonly used model of liver cell biology and express both SUN1 and lamin A/C; additionally, genotyping of this cell line determined that its endogenous *SUN1* alleles are wild-type for rs6461378 (Brian Halligan and Elizabeth Speliotes, *unpublished data*), making it an optimal system in which to test the functional effect of the H118Y variant of SUN1.

Huh7 cells were transfected with WT or H118Y SUN1 engineered to have a carboxy-terminal mCherry or myc-DDK tag in order to ensure that the tag would not interfere with the nucleoplasmic amino-terminus of SUN1. As expected, ectopically expressed tagged SUN1, either WT or H118Y, localized to the nuclear rim in transfected Huh7 cells, although the overall intensity of SUN1 staining was lower in H118Y-expressing cells (**Figure 3A**). Given that SUN1 was reported to be subject to proteasomal degradation in *Arabidopsis* (Huang et al., 2020), we hypothesized that one potential effect of the H118Y variant might be via differential regulation of protein turnover. When cells were maintained in complete medium containing 10% FBS, the H118Y variant did not appear to affect steady-state SUN1 levels (**Supplementary Figure 2**).

**Fig. 3.**
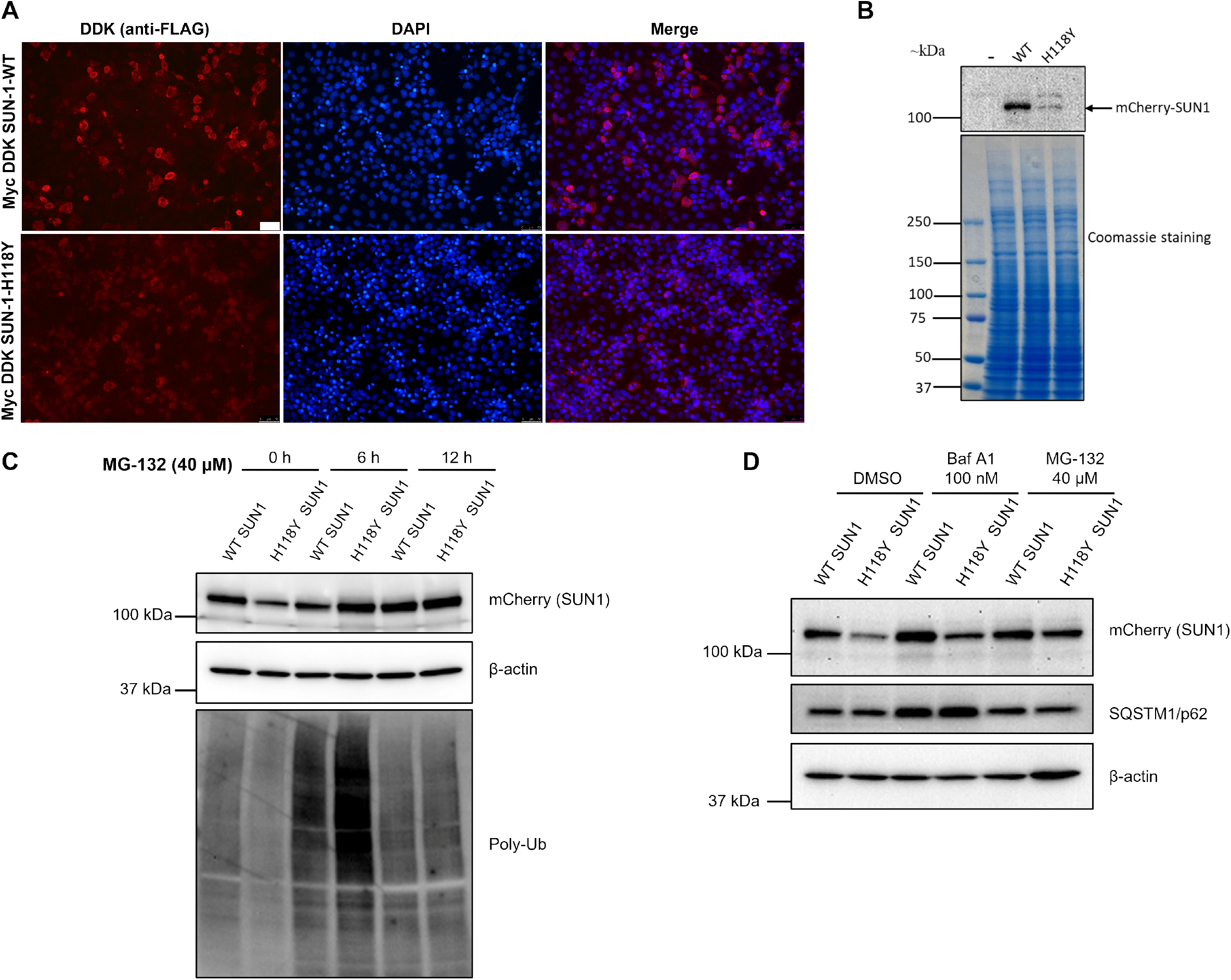
WT and H118Y SUN1 localize to the nuclear rim but undergo distinct cellular degradation pathways in Huh7 cells. (A) Localization of ectopically expressed SUN1 WT and H118Y was determined using anti-FLAG antibody (scale bar, 50 µm). (B) Immunoblot of mCherry-tagged SUN1 WT and SUN1 H118Y was performed using anti-mCherry antibody. Equal loading of samples was confirmed using Coomassie staining, and results shown were representative of n>5 independent experiments. (C) Immunoblot of SUN1 WT and H118Y after inhibiting proteasomal degradation; proteasomal inhibition was confirmed via anti-ubiquitin immunoblot. Representative images of n=3 independent experiments are shown. (D) Immunoblot of SUN1 WT and H118Y after inhibiting proteasomal or autophagic degradation pathway for 6 hours; inhibition of autophagy was confirmed via anti-SQSTM1/p62 immunoblot. Representative images of n=3 independent experiments are shown.

However, upon serum starvation of transfected cells for 48 hours, we consistently observed lower steady-state levels of SUN1 H118Y compared to WT as determined by immunoblot of whole cell lysates of Huh7 cells (**Figure 3B**). As expected, there was no change in *SUN1* transcript levels (**Supplementary Figure 3**), as *SUN1* transcription is governed by a CMV promoter in this expression construct. To address the possibility that the difference in steady-state SUN1 levels with serum starvation might be due to increased proteasomal degradation of SUN1 H118Y, transfected cells were treated with MG132 prior to lysis. Whereas the level of WT SUN1 was only modestly increased by MG132 treatment for up to 12 hours, the level of H118Y SUN1 was dramatically increased by MG132 and recovered to WT levels by 6 hours (**Figure 3C**). These data confirm for the first time in a mammalian system that SUN1 is subject to proteasomal degradation; furthermore, they indicate that the H118Y variant of SUN1 is subject to increased proteasomal degradation compared to WT SUN1. Given that we did not observe significant changes in WT SUN1 levels in the presence of MG132, and B-type lamins were previously reported to be degraded by autophagy (Dou et al., 2015), we hypothesized that degradation of WT SUN1 might occur primarily via autophagy. When Huh7 cells were transfected with WT or H118Y SUN1 and treated with MG132 or bafilomycin A1, we observed that bafilomycin A1 inhibited degradation of WT SUN1, with minimal impact on SUN1 H118Y, whereas MG132 again selectively blocked degradation of SUN1 H118Y (**Figure 3D**). These data are consistent with a model in which WT SUN1 is primarily degraded via autophagy in response to serum starvation, whereas the H118Y variant disrupts autophagic SUN1 degradation and is degraded by the proteasome. Furthermore, given that modulation of SUN1 levels has been shown to impact lamin A/C function and disease phenotypes in humans (Chai et al., 2021), this observation suggests a plausible mechanism by which rs6461378-T might contribute to metabolic disease.

### *The* SUN1 *H118Y variant increased insulin resistance and lipid accumulation in cells and is associated with increased expression of* NAFLD-*associated genes in human liver and Huh7 cellss*

Given that rs6461378-T associated positively with NAFLD and related metabolic traits in our discovery and validation cohorts, and that H118Y exhibited a biochemical phenotype in transfected cells, we reasoned that SUN1 H118Y might exhibit a direct metabolic effect in hepatocyte-like cells. Given that the strongest associations in our PheWAS analysis were with systemic metabolic traits including insulin resistance, we first asked whether expression of SUN1 H118Y would alter insulin sensitivity in transfected cells. Indeed, we found that expression of SUN1 H118Y decreased insulin sensitivity in Huh7 cells compared to either mock-transfected or WT SUN1-transfected cells, as determined by AKT phosphorylation after treatment with insulin (**Figure 4A**; quantitation of immunoblot data shown in **Figure 4B**). We observed a similar direct effect of SUN1 H118Y on lipid accumulation in Huh7 cells, as cells expressing SUN1 H118Y accumulated significantly more lipid than WT SUN1-expressing cells with or without oleic acid treatment (**Figure 4C**; quantitation shown in **Figure 4D**). Taken together, these data validate our genetic association data and suggest that the biochemical differences between WT and H118Y SUN1 lead to downstream functional consequences in cells, including insulin resistance and lipid accumulation.

**Fig. 4.**
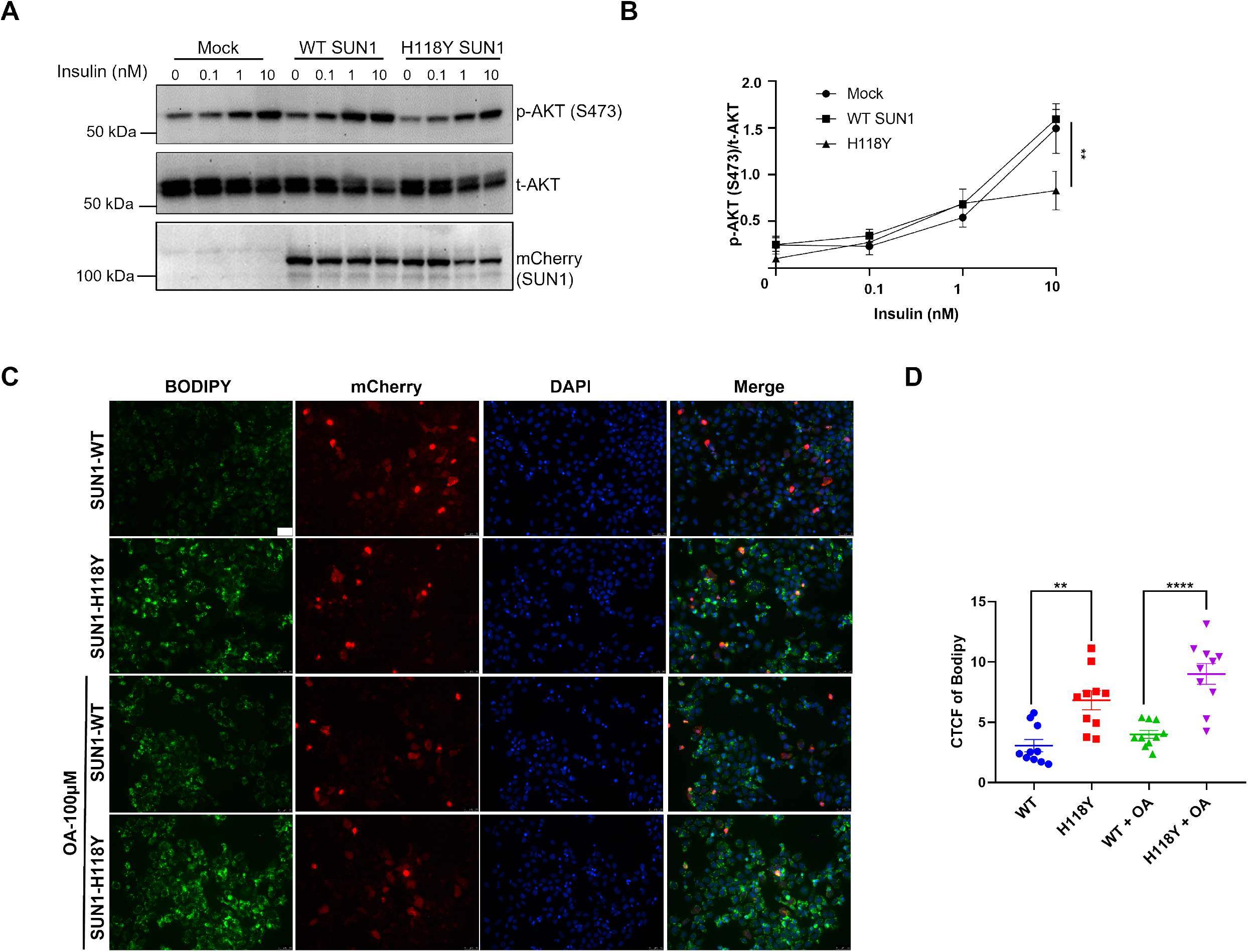
SUN1 H118Y impaired hepatic insulin signaling and increased lipid accumulation in Huh7 cells. (A) Representative immunoblot (n=3 independent experiments) showing phosphorylation of Ser-473 AKT after stimulation with the indicated insulin concentrations for 15 min. (B) Ser-473 phosphorylated AKT versus total AKT protein ratio was determined using ImageJ software, and dose-response curves were plotted in GraphPad Prism version 8; **, *P*<0.01 based on nonlinear regression analysis of dose-response curves (H118Y versus WT and mock-transfected). (C) Representative BODIPY 493/593 staining image from Huh7 cells, transfected with mCherry-tagged SUN1 WT and SUN1 H118Y plasmid for 24h and treated with oleic acid (100 µM) for an additional 24h; scale bar, 50 µm. (D) Lipid accumulation was quantified from n=3 independent experiments as the per-cell fluorescence of BODIPY 493/593 (CTCF). Data represented as mean ± S.E.M. \**P*<0.05, \*\**P*<0.01 or \*\*\**P*<0.001 relative to WT, as determined by one way or two way ANOVA followed by Tukey’s multiple comparison test.

The effects of rs6461378-T in human disease may be partly extra-hepatic, involving adipose tissue, skeletal muscle, small intestine, and/or the central nervous system in addition to the liver. Nevertheless, we hypothesized that if rs6461378-T predisposed to NAFLD, it would be positively associated with expression of NAFLD-related genes in the liver. To address this, we queried publicly available gene expression data via the GTEx Portal (GTEx version 8, http://gtexportal.org). We found that in liver tissue RNA isolated from the small cohort of genotyped individuals (n=208 in GTEx version 8), the presence of rs6461378-T was positively associated with hepatic expression of *SREBF1* (*P*=0.0036; **Figure 5A**), which is strongly linked to insulin resistance and lipogenesis (Shimomura et al., 2000). Positive associations with hepatic expression of *CD36* and *ACACA*, which are also linked to lipid uptake and lipogenesis (Hajri et al., 2002; Koonen et al., 2007), were seen as well, though these latter associations did not reach statistical significance. There was a parallel decrease in hepatic *GCKR* expression. Notably, there was also a subtle but notable increase in expression of *UBD* (*P*=0.07) and *CX3CR1* (*P*<0.05), which are associated with NASH, Mallory-Denk body formation, and progression of fibrosis (French et al., 2012; Wasmuth et al., 2008), among carriers of rs6461378-T. Notably, the power to detect differences in hepatic gene expression was limited in this small cohort of participants for whom liver transcriptome data and rs6461378 genotype are available; therefore, detection of differential expression of these NAFLD/NASH-related genes supports a causal link between rs6461378 and NAFLD and associated traits in humans. We corroborated these results by overexpressing SUN1 WT and H118Y in Huh7 cells and assessing transcription of lipid-related genes by qPCR. Similar to what was seen in the human GTEx data, *CD36* was upregulated in Huh7 cells expressing SUN1 H118Y compared to SUN1 WT (**Figure 5B**); additionally, while we did not observe a consistent change in *SREBF1* expression in this setting, there was upregulation of *ELOVL1* and *ELOVL5* (encoding fatty acid elongases), as well as *ANGPTL8*, a marker of insulin resistance (Chen et al., 2015), with downregulation of *ACOX1*, which encodes a key enzyme in the β-oxidation pathway. Taken together, these transcriptional data further support a causal relationship between rs6461378-T (*SUN1* H118Y) and hepatic steatosis, NASH, and metabolic disease in humans.

**Fig. 5.**
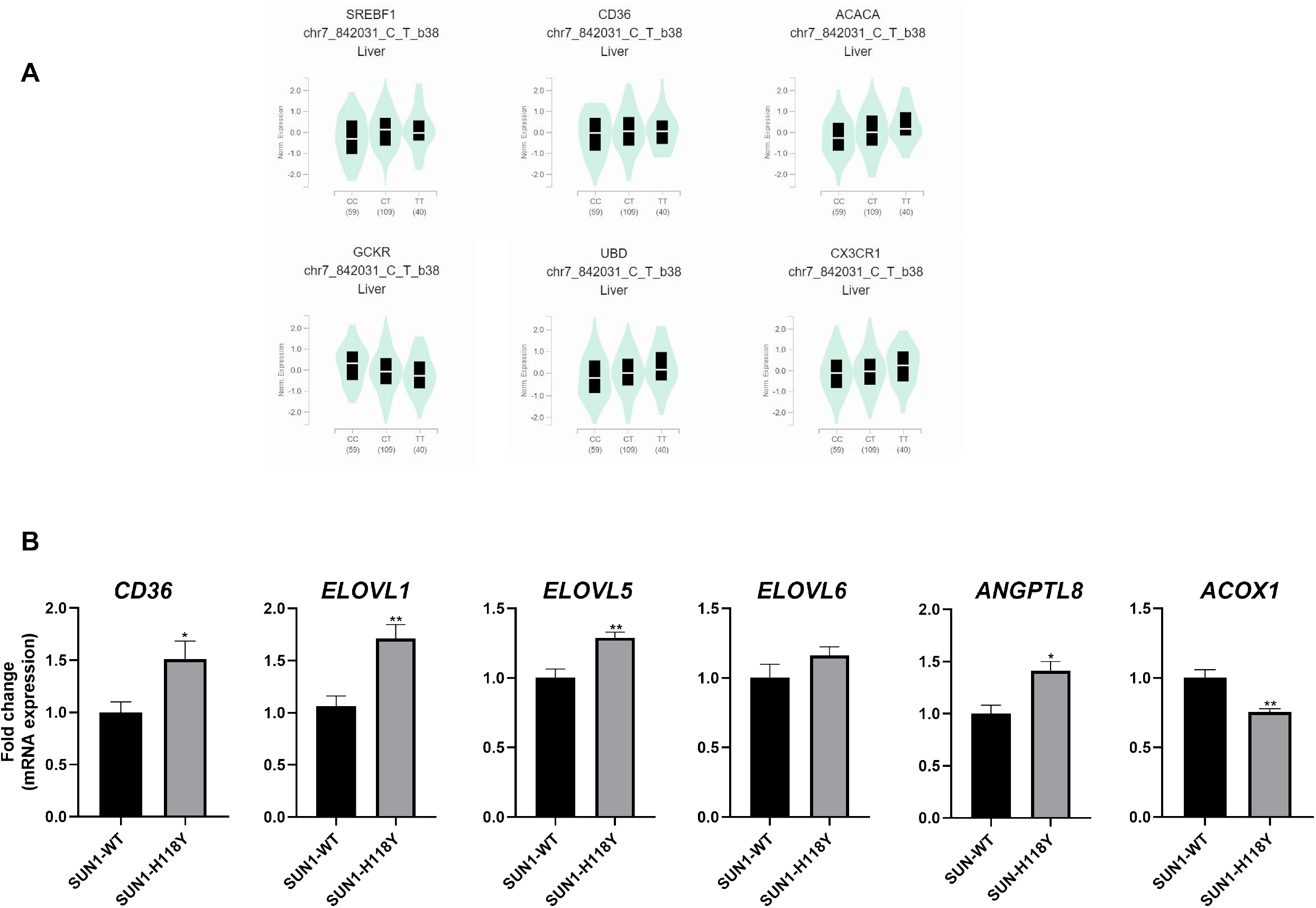
SUN1 H118Y alters transcription of lipid regulatory genes in human liver and Huh7 cells. (A) Violin plots of normalized transcript levels of the indicated genes in liver RNA in GTEx version 8 dataset for the indicated rs6461378 genotypes (CC, CT, TT) are shown. (B) Huh7 cells were transfected with mCherry-tagged SUN1 WT or H118Y plasmid followed by total RNA isolation after 48h, then qPCR to quantify transcript levels of the indicated genes related to fatty acid homeostasis (n=3 independent experiments). Data represented as mean ± S.E.M. \**P*<0.05, \*\**P*<0.01 or \*\*\**P*<0.001 relative to WT, as determined by Student’s t-test (two-tailed) with Mann-Whitney test.

## Discussion

Large-scale GWAS studies have identified several NAFLD susceptibility loci (Anstee et al., 2016; Speliotes et al., 2011), though these loci cannot account for the high degree of heritability suggested by twin and familial aggregation studies (Anstee et al., 2016; Cui et al., 2016; Grove et al., 2016; Loomba et al., 2015). Given that a subset of patients with mutations in *LMNA* develop lipodystrophy, metabolic syndrome, and NAFLD (Ajluni et al., 2017; Guenantin et al., 2014), we reasoned that other variants in genes encoding lamin-related and lamina-associated proteins might be found in individuals with NAFLD. Our meta-analysis of three large GWAS datasets identified a common coding variant within *SUN1* that positively associated with hepatic steatosis in all cohorts. Furthermore, this variant associated with multiple related metabolic traits in ancestrally distinct validation cohorts. To our knowledge, these are the first data linking a common nuclear envelope-related coding variant to NAFLD and metabolic disease.

A functional role for rs6461378-T was supported by *in silico* analysis, and testing in cell culture determined that this variant led to a biochemical phenotype, causing increased proteasomal degradation of SUN1 in response to serum starvation. Furthermore, while WT SUN1 was degraded primarily via autophagy, SUN1 H118Y was resistant to autophagic degradation, and expression of SUN1 H118Y decreased insulin sensitivity and increased lipid accumulation in Huh7 cells. These results align with previous studies in mice indicating a role for altered SUN1 degradation in laminopathies (Chen et al., 2012). Additionally, these data were consistent with publicly available gene expression data in GTEx version 8, which demonstrated that rs6461378-T was associated with increased hepatic expression of *SREBF1* and other lipid-related and pro -inflammatory genes. Collectively, these data suggest that this variant may have a direct functional role in human disease.

While rare autosomal dominant mutations in *LMNA* have been recognized as a cause of genetically determined lipodystrophy syndromes for more than two decades, common variants in nuclear envelope and nuclear lamina-related genes that contribute to metabolic disease and NASH at a population level have been elusive. Here we provide the first evidence from multiple large cohorts, including large and ethnically distinct validation cohorts, for a potential pathogenic role of a coding variant in *SUN1*. The characterization of this *SUN1* variant and its role in metabolic disease has the potential to inform our understanding of the pathogenesis of laminopathy syndromes and the roles of genetic modifiers in determining their penetrance and severity. In addition, the identification of this common *SUN1* variant associated with NASH, insulin resistance, and metabolic disease puts the nuclear envelope squarely among the subcellular structures with important roles in the pathogenesis of these diseases. Prior work from our group has suggested based on pre-clinical studies that a lamin-associated protein, LAP2α, might be a promising therapeutic target to enhance the hepatoprotective effects of lamin A/C in NASH (Upadhyay et al., 2022); now, this study suggests that SUN1 might also be a potential target, with a common coding variant that may play a functional role in a significant proportion of patients with NASH. This is of particular relevance because it appears quite likely, based on the clinical trial evidence available to date, that combinatorial and patient-targeted therapeutic approaches may be increasingly utilized in the treatment of NASH (Alkhouri et al., 2022).

It is notable that rs6461378-T, which has strong positive associations with hepatic steatosis and NASH-related traits and a biochemical phenotype in cells, is quite common in the population, despite SUN1 His118 being a conserved residue in mammals (Supplementary Figure 1). Furthermore, available data from population-based studies suggest that its allele frequency varies significantly among racial and ethnic groups, and in some populations it may even be the major allele (Phan et al., 2020). While perhaps surprising, this could reflect changes in energy storage-related evolutionary pressures over time, as has been proposed for common risk alleles with strong effects such as rs738409 (*PNPLA3* I148M; Carlsson et al., 2020). Moreover, the frequency of rs6461378-T underscores its potential importance, as at least half of individuals with NASH are predicted to carry one or more rs6461378-T alleles.

Our study has some important limitations. First, due to our requirement that genotype data be available from at least two of three discovery cohorts in order to include a variant in the association meta-analysis, many coding variants in *SUN1* and other nuclear envelope-related genes were excluded from our analyses. Thus, additional genetic associations may exist that could not be detected in this study. Furthermore, although our corroborative findings in large validation cohorts, *in silico* analysis, and cell culture studies are supportive of a causal role for rs6461378-T in NASH and related metabolic traits, the association analyses in this study cannot establish a definite causal relationship. Strengths of this study include the multiple large, well-phenotyped and validated discovery cohorts, the stringency of inclusion criteria for coding variants, the large and ethnically distinct validation cohorts, a plausible biological mechanism by which SUN1 H118Y could contribute to the pathogenesis of NASH and metabolic disease, and cell culture data demonstrating a biochemical phenotype consistent with such a mechanism.

In summary, here we provide the first evidence of a genetic link between a common nuclear envelope-related coding variant and NASH and related traits at a population level. Moreover, our biochemical and functional validation studies suggest that SUN1 may be a promising and relevant therapeutic target in NASH and metabolic disease, particularly given how commonly this variant is found in the population. Thus, it will be critical to establish via relevant disease models whether the relationship between rs6461378-T and metabolic disease is truly causal, and whether this is determined by altered degradation of the encoded protein. Future genetic association studies should explore whether additional variants in *SUN1*, and related nuclear envelope-encoding genes, might also play causal roles in NASH and metabolic disease. As noted above, our study excluded variants for which data were not present in at least two of the discovery cohorts, and thus additional genetic associations could have gone undetected. Finally, this study suggests an important relationship between inner nuclear membrane protein degradation and cellular metabolism that may have broad relevance to human health. A full understanding of the mediators and mechanisms by which nuclear membrane proteins are degraded will be critical in order to maximize its potential yield as a therapeutic target in NASH, metabolic disease, and beyond.

## Data Availability

All data produced in the present study are contained in the manuscript or available upon reasonable request to the authors.

## Acknowledgement

The authors would like to thank members of the Brady and Speliotes laboratories for helpful discussions and Brian Halligan, PhD, for sharing unpublished Huh7 *SUN1* genotyping data. In addition, the authors are grateful to Brandon Buscher, BS, for technical assistance.

## References

Ajluni, N., Meral, R., Neidert, A.H., Brady, G.F., Buras, E., McKenna, B., DiPaola, F., Chenevert, T.L., Horowitz, J.F., Buggs-Saxton, C., et al. (2017). Spectrum of disease associated with partial lipodystrophy: lessons from a trial cohort. Clin Endocrinol (Oxf) 86, 698–707.

Alkhouri, N., Herring, R., Kabler, H., Kayali, Z., Hassanein, T., Kohli, A., Huss, R.S., Zhu, Y., Billin, A.N., Damgaard, L.H., et al. (2022). Safety and efficacy of combination therapy with semaglutide, cilofexor and firsocostat in patients with non-alcoholic steatohepatitis: A randomised, open-label phase II trial. J Hepatol 77, 607–618.

Anstee, Q.M., Seth, D., and Day, C.P. (2016). Genetic Factors That Affect Risk of Alcoholic and Nonalcoholic Fatty Liver Disease. Gastroenterology 150, 1728–1744 e1727.

Axelsen, M., Smith, U., Eriksson, J.W., Taskinen, M.R., and Jansson, P.A. (1999). Postprandial hypertriglyceridemia and insulin resistance in normoglycemic first-degree relatives of patients with type 2 diabetes. Ann Intern Med 131, 27–31.

Brady, G.F., Kwan, R., Bragazzi Cunha, J., Elenbaas, J.S., and Omary, M.B. (2018a). Lamins and lamin-associated proteins in gastrointestinal health and disease. Gastroenterology 154, 1602–1619.

Brady, G.F., Kwan, R., Ulintz, P.J., Nguyen, P., Bassirian, S., Basrur, V., Nesvizhskii, A.I., Loomba, R., and Omary, M.B. (2018b). Nuclear lamina genetic variants, including a truncated LAP2, in twins and siblings with nonalcoholic fatty liver disease. Hepatology 67, 1710–1725.

Carlsson, B., Linden, D., Brolen, G., Liljeblad, M., Bjursell, M., Romeo, S., and Loomba, R. (2020). Review article: the emerging role of genetics in precision medicine for patients with non-alcoholic steatohepatitis. Aliment Pharmacol Ther 51, 1305–1320.

Chai, R.J., Werner, H., Li, P.Y., Lee, Y.L., Nyein, K.T., Solovei, I., Luu, T.D.A., Sharma, B., Navasankari, R., Maric, M., et al. (2021). Disrupting the LINC complex by AAV mediated gene transduction prevents progression of Lamin induced cardiomyopathy. Nat Commun 12, 4722.

Chen, C.Y., Chi, Y.H., Mutalif, R.A., Starost, M.F., Myers, T.G., Anderson, S.A., Stewart, C.L., and Jeang, K.T. (2012). Accumulation of the inner nuclear envelope protein Sun1 is pathogenic in progeric and dystrophic laminopathies. Cell 149, 565–577.

Chen, X., Lu, P., He, W., Zhang, J., Liu, L., Yang, Y., Liu, Z., Xie, J., Shao, S., Du, T., et al. (2015). Circulating betatrophin levels are increased in patients with type 2 diabetes and associated with insulin resistance. J Clin Endocrinol Metab 100, E96–100.

Chen, V.L., Du, X., Chen, Y., Kuppa, A., Handelman, S.K., Vohnoutka, R.B., Peyser, P.A., Palmer, N.D., Bielak, L.F., Halligan, B., et al. (2021). Genome-wide association study of serum liver enzymes implicates diverse metabolic and liver pathology. Nat Commun 12, 816.

Cotter, T.G., and Rinella, M. (2020). Nonalcoholic Fatty Liver Disease 2020: The State of the Disease. Gastroenterology 158, 1851–1864.

Cui, J., Chen, C.H., Lo, M.T., Schork, N., Bettencourt, R., Gonzalez, M.P., Bhatt, A., Hooker, J., Shaffer, K., Nelson, K.E., et al. (2016). Shared genetic effects between hepatic steatosis and fibrosis: A prospective twin study. Hepatology 64, 1547–1558.

Dou, Z., Xu, C., Donahue, G., Shimi, T., Pan, J.A., Zhu, J., Ivanov, A., Capell, B.C., Drake, A.M., Shah, P.P., et al. (2015). Autophagy mediates degradation of nuclear lamina. Nature 527, 105–109.

French, S.W., French, B.A., Oliva, J., Li, J., Bardag-Gorce, F., Tillman, B., and Canaan, A. (2012). FAT10 knock out mice livers fail to develop Mallory-Denk bodies in the DDC mouse model. Exp Mol Pathol 93, 309–314.

Graumann, K., Runions, J., and Evans, D.E. (2010). Characterization of SUN-domain proteins at the higher plant nuclear envelope. Plant J 61, 134–144.

Grove, J.I., Austin, M., Tibble, J., Aithal, G.P., and Verma, S. (2016). Monozygotic twins with NASH cirrhosis: cumulative effect of multiple single nucleotide polymorphisms? Ann Hepatol 15, 277–282.

Guenantin, A.C., Briand, N., Bidault, G., Afonso, P., Bereziat, V., Vatier, C., Lascols, O., Caron-Debarle, M., Capeau, J., and Vigouroux, C. (2014). Nuclear envelope-related lipodystrophies. Semin Cell Dev Biol 29, 148–157.

Hajri, T., Han, X.X., Bonen, A., and Abumrad, N.A. (2002). Defective fatty acid uptake modulates insulin responsiveness and metabolic responses to diet in CD36-null mice. J Clin Invest 109, 1381–1389.

Haque, F., Lloyd, D.J., Smallwood, D.T., Dent, C.L., Shanahan, C.M., Fry, A.M., Trembath, R.C., and Shackleton, S. (2006). SUN1 interacts with nuclear lamin A and cytoplasmic nesprins to provide a physical connection between the nuclear lamina and the cytoskeleton. Mol Cell Biol 26, 3738–3751.

Huang, A., Tang, Y., Shi, X., Jia, M., Zhu, J., Yan, X., Chen, H., and Gu, Y. (2020). Proximity labeling proteomics reveals critical regulators for inner nuclear membrane protein degradation in plants. Nat Commun 11, 3284.

Konerman, M.A., Jones, J.C., and Harrison, S.A. (2018). Pharmacotherapy for NASH: Current and emerging. J Hepatol 68, 362–375.

Koonen, D.P., Jacobs, R.L., Febbraio, M., Young, M.E., Soltys, C.L., Ong, H., Vance, D.E., and Dyck, J.R. (2007). Increased hepatic CD36 expression contributes to dyslipidemia associated with diet-induced obesity. Diabetes 56, 2863–2871.

Kwan, R., Brady, G.F., Brzozowski, M., Weerasinghe, S.V., Martin, H., Park, M.J., Brunt, M.J., Menon, R.K., Tong, X., Yin, L., et al. (2017). Hepatocyte-Specific Deletion of Mouse Lamin A/C Leads to Male-Selective Steatohepatitis. Cell Mol Gastroenterol Hepatol 4, 365–383.

Le Dour, C., Wu, W., Bereziat, V., Capeau, J., Vigouroux, C., and Worman, H.J. (2017). Extracellular matrix remodeling and transforming growth factor-beta signaling abnormalities induced by lamin A/C variants that cause lipodystrophy. J Lipid Res 58, 151–163.

Locke, A.E., Steinberg, K.M., Chiang, C.W.K., Service, S.K., Havulinna, A.S., Stell, L., Pirinen, M., Abel, H.J., Chiang, C.C., Fulton, R.S., et al. (2019). Exome sequencing of Finnish isolates enhances rare-variant association power. Nature 572, 323–328.

Loomba, R., Schork, N., Chen, C.H., Bettencourt, R., Bhatt, A., Ang, B., Nguyen, P., Hernandez, C., Richards, L., Salotti, J., et al. (2015). Heritability of Hepatic Fibrosis and Steatosis Based on a Prospective Twin Study. Gastroenterology 149, 1784–1793.

Paik, J.M., Golabi, P., Biswas, R., Alqahtani, S., Venkatesan, C., and Younossi, Z.M. (2020). Nonalcoholic Fatty Liver Disease and Alcoholic Liver Disease are Major Drivers of Liver Mortality in the United States. Hepatol Commun 4, 890–903.

Palmer, N.D., Kahali, B., Kuppa, A., Chen, Y., Du, X., Feitosa, M.F., Bielak, L.F., O’Connell, J.R., Musani, S.K., Guo, X., et al. (2021). Allele-specific variation at APOE increases nonalcoholic fatty liver disease and obesity but decreases risk of Alzheimer’s disease and myocardial infarction. Hum Mol Genet 30, 1443–1456.

Phan, L., Jin, Y., Zhang, H., Qiang, W., Shekhtman, E., Shao, D., Revoe, D., Villamarin, R., Ivanchenko, E., Kimura, M., et al. (2020). ALFA: Allele Frequency Aggregator. National Center for Biotechnology Information, U.S. National Library of Medicine, 10 Mar. 2020, www.ncbi.nlm.nih.gov/snp/docs/gsr/alfa/.

Shackleton, S., Lloyd, D.J., Jackson, S.N., Evans, R., Niermeijer, M.F., Singh, B.M., Schmidt, H., Brabant, G., Kumar, S., Durrington, P.N., et al. (2000). LMNA, encoding lamin A/C, is mutated in partial lipodystrophy. Nat Genet 24, 153–156.

Shimomura, I., Matsuda, M., Hammer, R.E., Bashmakov, Y., Brown, M.S., and Goldstein, J.L. (2000). Decreased IRS-2 and increased SREBP-1c lead to mixed insulin resistance and sensitivity in livers of lipodystrophic and ob/ob mice. Mol Cell 6, 77–86.

Shin, J.Y., Hernandez-Ono, A., Fedotova, T., Ostlund, C., Lee, M.J., Gibeley, S.B., Liang, C.C., Dauer, W.T., Ginsberg, H.N., and Worman, H.J. (2019). Nuclear envelope-localized torsinA-LAP1 complex regulates hepatic VLDL secretion and steatosis. J Clin Invest 129, 4885–4900.

Speliotes, E.K., Yerges-Armstrong, L.M., Wu, J., Hernaez, R., Kim, L.J., Palmer, C.D., Gudnason, V., Eiriksdottir, G., Garcia, M.E., Launer, L.J., et al. (2011). Genome-wide association analysis identifies variants associated with nonalcoholic fatty liver disease that have distinct effects on metabolic traits. PLoS Genet 7, e1001324.

Upadhyay, K.K., Choi, E.-Y.K., Foisner, R., Omary, M.B., and Brady, G.F. (2022). Hepatocyte-specific loss of LAP2α protects against diet-induced hepatic steatosis and steatohepatitis in male mice. bioRxiv 484917 [Preprint]. March 19, 2022; doi: 10.1101/2022.03.18.484917.

Wang, Y., Meng, X., Deng, X., Okekunle, A.P., Wang, P., Zhang, Q., Ding, L., Guo, X., Lv, M., Sun, C., et al. (2018). Postprandial Saturated Fatty Acids Increase the Risk of Type 2 Diabetes: A Cohort Study in a Chinese Population. J Clin Endocrinol Metab 103, 1438–1446.

Wasmuth, H.E., Zaldivar, M.M., Berres, M.L., Werth, A., Scholten, D., Hillebrandt, S., Tacke, F., Schmitz, P., Dahl, E., Wiederholt, T., et al. (2008). The fractalkine receptor CX3CR1 is involved in liver fibrosis due to chronic hepatitis C infection. J Hepatol 48, 208–215.

Wojtanik, K.M., Edgemon, K., Viswanadha, S., Lindsey, B., Haluzik, M., Chen, W., Poy, G., Reitman, M., and Londos, C. (2009). The role of LMNA in adipose: a novel mouse model of lipodystrophy based on the Dunnigan-type familial partial lipodystrophy mutation. J Lipid Res 50, 1068–1079.

Wong, R.J., Aguilar, M., Cheung, R., Perumpail, R.B., Harrison, S.A., Younossi, Z.M., and Ahmed, A. (2015). Nonalcoholic steatohepatitis is the second leading etiology of liver disease among adults awaiting liver transplantation in the United States. Gastroenterology 148, 547–555.

Younossi, Z.M., Loomba, R., Rinella, M.E., Bugianesi, E., Marchesini, G., Neuschwander-Tetri, B.A., Serfaty, L., Negro, F., Caldwell, S.H., Ratziu, V., et al. (2018). Current and future therapeutic regimens for nonalcoholic fatty liver disease and nonalcoholic steatohepatitis. Hepatology 68, 361–371.

