## Supplementary Material for "A common variant that alters SUN1 degradation associates with hepatic steatosis and metabolic traits in multiple cohorts"

### Supplementary Fig 1

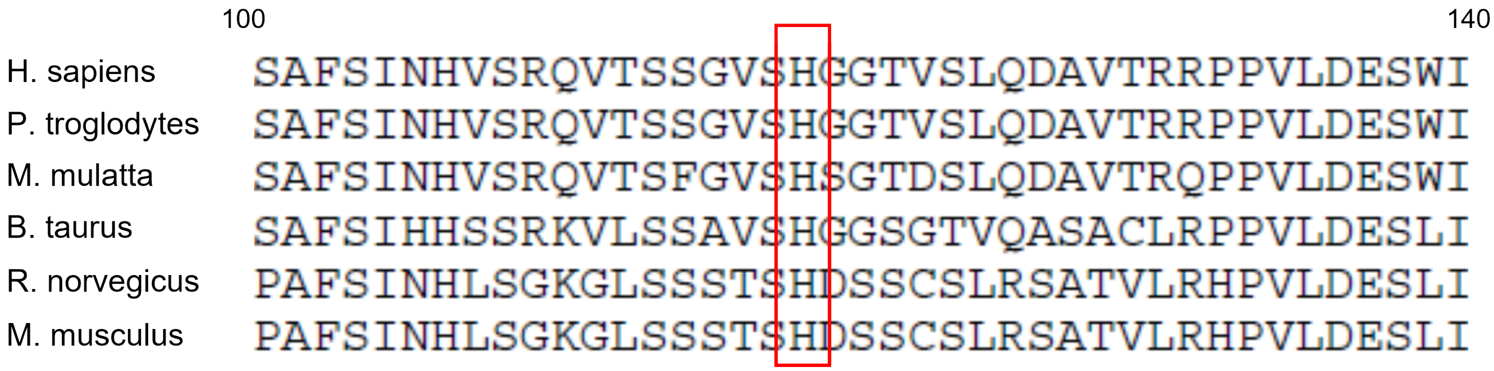


**Supplementary Fig.1. Alignment of homologous mammalian SUN1 amino acid sequences.** Alignment of selected mammalian SUN1 amino acid sequences in the region of His118 was performed using the web-based tool Benchling (http://benchling.com). Codon number labels refer to the human SUN1 sequence; His118 is highlighted in the red box.

#### Supplementary Fig 2

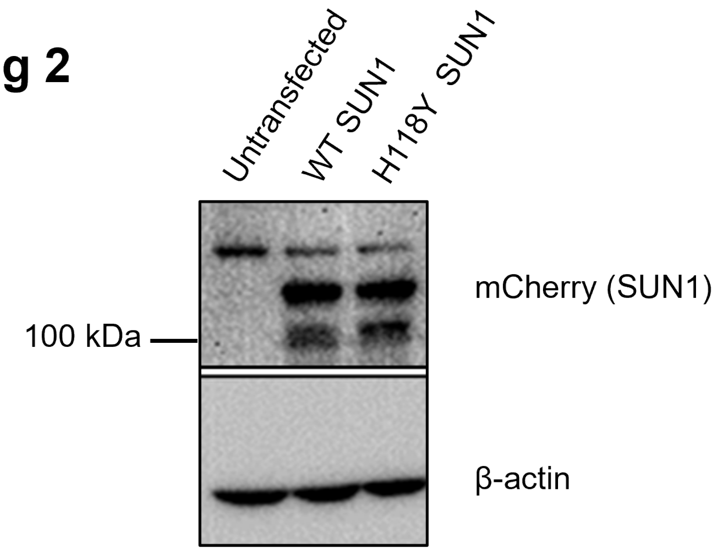


**Supplementary Fig.2. Equivalent expression of SUN1 WT and H118Y in the presence of complete medium.** Huh7 cells were transfected with mCherry-tagged SUN1 WT or H118Y and grown in complete medium containing 10% FBS for 48h prior to lysis, SDS-PAGE, and immunoblot using anti-mCherry antibody. Equal loading of samples was confirmed using β-actin immunoblot, and results shown were representative of n=3 independent experiments.

### Supplementary Fig 3

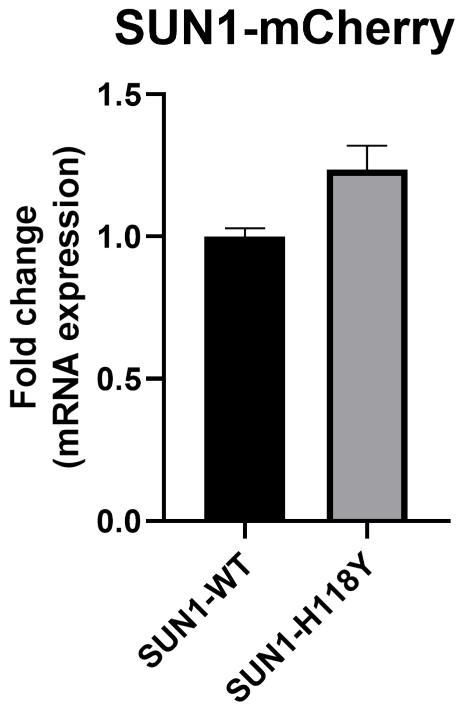


**Supplementary Fig.3. SUN1-mCherry transcript levels do not differ between SUN1 WT and H118Y-transfected cells.** Huh7 cells were transfected with mCherry tagged SUN1 WT and SUN1 H118Y; after 48h, total RNA was isolated, and SUN1-mCherry transcript levels were determined via qPCR. Data were derived from n=3 independent experiments and are represented as mean ± S.E.M.

| Target | Catalog Number & manufacturer | Dilution |
|---|---|---|
| Phospho-Akt (Ser473) | 4060; Cell Signaling | 1:1000 (IB) |
| Phospho-Akt (T-308) | 13038; Cell Signaling | 1:1000 (IB) |
| AKT (total) | 4691; Cell Signaling | 1:1000 (IB) |
| mCherry | PA5-34974; Invitrogen | 1:1000 (IB) |
| FLAG/DDK tag | F3165; Sigma | 1:400 (IF) |
| SQSTM1/p62 | 5114; Cell Signaling | 1:1000 (IB) |
| Ubiquitin | 14-6078-80; Invitrogen | 1:250 (IB) |
| β-actin | 4970, Cell Signaling | 1:1500 (IB) |
| Anti-Rabbit IgG-HRP | A6154; Sigma | 1:1500 (IB) |
| Anti-Mouse IgG-HRP | 32430; Invitrogen | 1:1500 (IB) |
| Goat anti-Mouse IgG, Alexa Fluor™ 680 | A21057; Invitrogen | 1:400 (IF) |

**Supplementary Table 1.** Antibodies and dilutions used (IB: Immunoblot, IF: Immunofluorescence).

| Gene | Forward (5′→3′) | Reverse (5′→3′) |
| --- | --- | --- |
| *CD36* | CAG GTC AACCTA TTG GTC AAG CC | GCC TTC TCA TCACCA ATG GTC C |
| *ELOVL1* | GTC TAC AACTTC TCA CTG GTG GC | AAG TGC CTCAGG GCT GTT GGA A |
| *ELOVL5* | ACG TCT ACCACC ATG CCT CGA | TGG AAG GGACTG ACG ACA AAC C |
| *ELOVL6* | CCA TCC AATGGA TGC AGG AAA AC | CCA GAG CACTAA TGG CTT CCT C |
| *ANGPTL8* | GCAAGCCTGTTGGAGACTCAGA | GCTCCTCAGCTGGACTTCTAGC |
| *ACOX1* | GGC GCA TAC ATG AAG GAG ACC T | AGG TGA AAGCCT TCA GTC CAG C |
| *18S* | ACCCGTTGAACCCCATTCGTGA | GCCTCACTAAACCATCCAATCGG |
| *SUN1-mCherry* | GAG GCT GAA GCT GAA GGA C | GAT GGT GTA GTC CTC GTT GTG |

**Supplementary Table 2.** Primers used in qPCR analysis.
